# Research ethics review during the COVID-19 pandemic: An international study

**DOI:** 10.1101/2023.09.24.23296056

**Authors:** Fabio Salamanca-Buentello, Rachel Katz, Diego S. Silva, Ross E.G. Upshur, Maxwell J. Smith

## Abstract

Research ethics review committees (ERCs) worldwide faced daunting challenges during the COVID-19 pandemic. There was a need to balance rapid turnaround with rigorous evaluation of high-risk research protocols in the context of considerable uncertainty. This study explored the experiences and performance of ERCs during the pandemic.

We conducted an anonymous, cross-sectional, global online survey of chairs (or their delegates) of ERCs who were involved in the review of COVID-19-related research protocols after March 2020. The survey ran from October 2022 to February 2023 and consisted of 50 items, with opportunities for open text responses.

Two hundred and three participants [130 from high-income countries (HICs) and 73 from low-and middle-income countries (LMICs)] completed our survey. Respondents came from diverse entities and organizations from 48 countries (19 HICs and 29 LMICs) in all World Health Organization regions. Responses show little of the increased global funding for COVID-19 research was allotted to the operation of ERCs. Few ERCs had pre-existing internal policies to address operation during public health emergencies, but almost half used existing guidelines. Most ERCs modified existing procedures or designed and implemented new ones but had not evaluated the success of these changes. Participants overwhelmingly endorsed permanently implementing several of them. Few ERCs added new members but non-member experts were consulted; quorum was generally achieved. Collaboration among ERCs was infrequent, but reviews conducted by external ERCs were recognized and validated. Review volume increased during the pandemic, with COVID-19-related studies being prioritized. Most protocol reviews were reported as taking less than three weeks. One-third of respondents reported external pressure on their ERCs from different stakeholders to approve or reject specific COVID-19-related protocols.

ERC members faced significant challenges to keep their committees functioning during the pandemic. Our findings can inform ERC approaches towards future public health emergencies. To our knowledge, this is the first international, COVID-19-related study of its kind.

## INTRODUCTION

The ethical review of research protocols during public health emergencies (PHEs) such as the COVID-19 pandemic is a daunting task. Committees tasked with assessing the ethical acceptability of research projects, which we refer to as ethics review committees (ERCs) but are also variably called research ethics boards, research ethics committees, ethics review boards, and institutional review boards, face challenges to reviewing research protocols swiftly while maintaining a high degree of rigour, all under suboptimal conditions and uncertainty. ERCs must balance the urge for rapid turnaround and flexibility with the requirement for intense scrutiny given that new projects often propose innovative but high-risk diagnostic, therapeutic, or preventive approaches to address the PHE. This is especially challenging in the case of countries with fragile health systems, poor infrastructure, and little experience conducting medical research, as well as countries experiencing protracted emergencies [1–5].

Failure to ensure rigour and depth during rapid ethics reviews in public health emergencies may place research participants at risk [6]. In such challenging circumstances, ERCs must consider how interventions, study design, eligibility criteria, community engagement, and approaches to vulnerable populations impact scientific validity, participant autonomy, respect for persons, welfare, justice, and social value [2,7–9]. Additional demands on ERCs may include the ability to incorporate and respond swiftly to newly available knowledge, provide monitoring and oversight of research, and considerations of the impact of the PHE on those involved in the research process, such as research participants, investigators, and ERC members and staff [7].

Public health emergencies force ERCs to make reasonable adjustments and design innovative strategies to address the various components of research ethics review while still adhering to ethical principles [3,6,7,10]. Moreover, after a PHE, changes implemented to secure continued operations of ERCs must be evaluated to determine their success and whether they should be permanently put in place to improve the everyday functioning of the committees.

Given the challenges that ERCs worldwide have faced during the COVID-19 pandemic, we aimed in this exploratory study to identify their experiences in the attempt to adapt to this PHE. We were particularly interested in the availability of pandemic-specific support, the promptness of protocol review, the volume of protocols received, the modifications to and innovations in operational procedures and policies and the evaluation of their outcomes, the anticipated permanence of such changes beyond the pandemic, the presence of pressure from different stakeholders on ERCs, the efforts to ensure quorum, the changes to the composition of ERCs, and the approaches to strengthen inter-ERC collaboration. To our knowledge, this is the first international, COVID-19-related study of its kind.

## METHODS

This international, cross-sectional, exploratory online survey was conducted by researchers from Western University, the University of Toronto, and the Lunenfeld – Tanenbaum Research Institute in Canada, and the University of Sydney in Australia, in collaboration with the World Health Organization’s COVID-19 Ethics and Governance Working Group.

### Inclusion criteria

We used targeted purposive and criterion sampling to invite Chairs and members of ERCs who were actively involved in the ethics review of COVID-19 research protocols to participate in this study. To ensure eligibility of participants, the first question of the survey asked respondents to confirm whether they had reviewed COVID-19-related research protocols during the pandemic. Responding to our survey was entirely voluntary. For the purposes of this study, we considered March 2020 as the beginning of this PHE. We specifically targeted individuals from all WHO regions. Participants were assigned to either of two categories: high-income countries (HICs) or low-and middle-income countries (LMICs), according to their reported country of residence. To do this, we used the World Bank classification of countries (https://datahelpdesk.worldbank.org/knowledgebase/articles/906519-world-bank-country-and-lending-groups), which is based on gross national income per capita. We adopted this widely-used categorization notwithstanding its limitations in terms of hiding power imbalances and reducing important differences to questions of economics [11].

### Survey questionnaire

The complete questionnaire is available as **S1 Appendix**. The overall structure and flow of the survey questionnaire, which consisted of a main “trunk” of 37 items organized into 11 thematic categories, is shown in **Error! Reference source not found.**. Eight of these items branched into different survey flow elements based on respondents’ answers. Thus, in total, the questionnaire, written in English, included 50 questions. We privileged close-ended over open-ended questions, but we allowed respondents the opportunity to provide additional comments for some items. We pilot-tested the online questionnaire with a small group of experts who fulfilled the inclusion criteria. This helped polish the wording of the questions and also assess and improve the logistics of the administration of the survey.

### Data collection

The invitation to participate in the survey explained the nature and purpose of our study, the inclusion criteria used to select participants, a summary of the procedures involved, and the URL link to the survey. These invitations were initially distributed by email by the WHO’s COVID-19 Ethics and Governance Working Group through the email listserv of the 13th Global Summit of National Ethics Committees (an event that took place in September 2022). The Working Group identified additional potential participants among its extensive contact networks. We also circulated the invitation to experts identified by the research team. Invitations could also be forwarded to individuals designated by ERCs. In both cases, those invited fulfilled our inclusion criteria.

Our survey was active from October 11, 2022, to February 28, 2023. We used the Qualtrics Experience Management (XM) online platform to administer the questionnaire, which was open only to individuals who received the invitation with the link to the survey.

### Data analysis

The analysis of the findings of this exploratory study employed descriptive statistics and stratified the comparison between responses of participants from HICs with those of participants from LMICs. To facilitate the examination of the results, tables were prepared showing the number and percentage of respondents from HICs and LMICs who answered each question in the survey. Qualitative data (text responses from open-ended questions) were evaluated using thematic analysis and the constant comparative method.

### Research ethics approval

Our study received approval from Western University’s Non-Medical Research Ethics Board (Protocol ID 120455). Additionally, it was evaluated by the World Health Organization Research Ethics Review Committee (Protocol ID CERC.0181) and was exempted from further review. The use of the Qualtrics platform facilitated data collection and management while respecting the privacy and confidentiality of participants. Respondents indicated their consent to participate in our survey by selecting a button labelled “I consent” at the end of the letter of information and consent, which appeared on the first page of the questionnaire. Responses were anonymous to protect participants’ privacy and confidentiality and encourage the open sharing of experiences.

## RESULTS

### Characterization of survey respondents

Two hundred and eighty-one individuals opened our survey. Of these, 250 answered the first question, which confirmed whether respondents fulfilled our inclusion criteria, and with which we could confirm their eligibility. Forty-three individuals explicitly indicated that they did not meet our criteria. Thus, the initial number of suitable respondents was 207. As expected in surveys such as ours in which participants are allowed to skip questions, the number of respondents per question varied slightly, from a maximum of 207 to a minimum of 147.

Of the 204 participants who indicated their sex / gender, 120 (58.8%) were female, 82 (40.2%) were male, one (0.5%) preferred to self-describe, and one (0.5%) preferred not to disclose this information (Box 1, Table a). The proportion of females was higher in HICs (64.9%) than in LMICs (47.9%); thus, the distribution of respondents by sex / gender was more balanced in LMICs than in HICs. As shown in Box 1, Table b, more than three quarters of respondents (77.9%) were 45 years old or older. This was true for both HICs and LMICs. Most respondents provided ethics review for national bodies, such as national ethics committees or national public health organizations; more than a quarter participated in ERCs linked to academic or research institutions (Box 1, Table c). However, while almost half of respondents from HICs were members of ERCs affiliated with national bodies, only one quarter of participants from LMICs provided ethics review for such organizations. In contrast, in LMICs, 40% of respondents were members of ERCs associated with academic or research institutions. Furthermore, only 20% of participants from LMICs and 13.9% of participants from HICs provided ethics review for health care facilities.

#### Box 1.

Characterization of survey participants

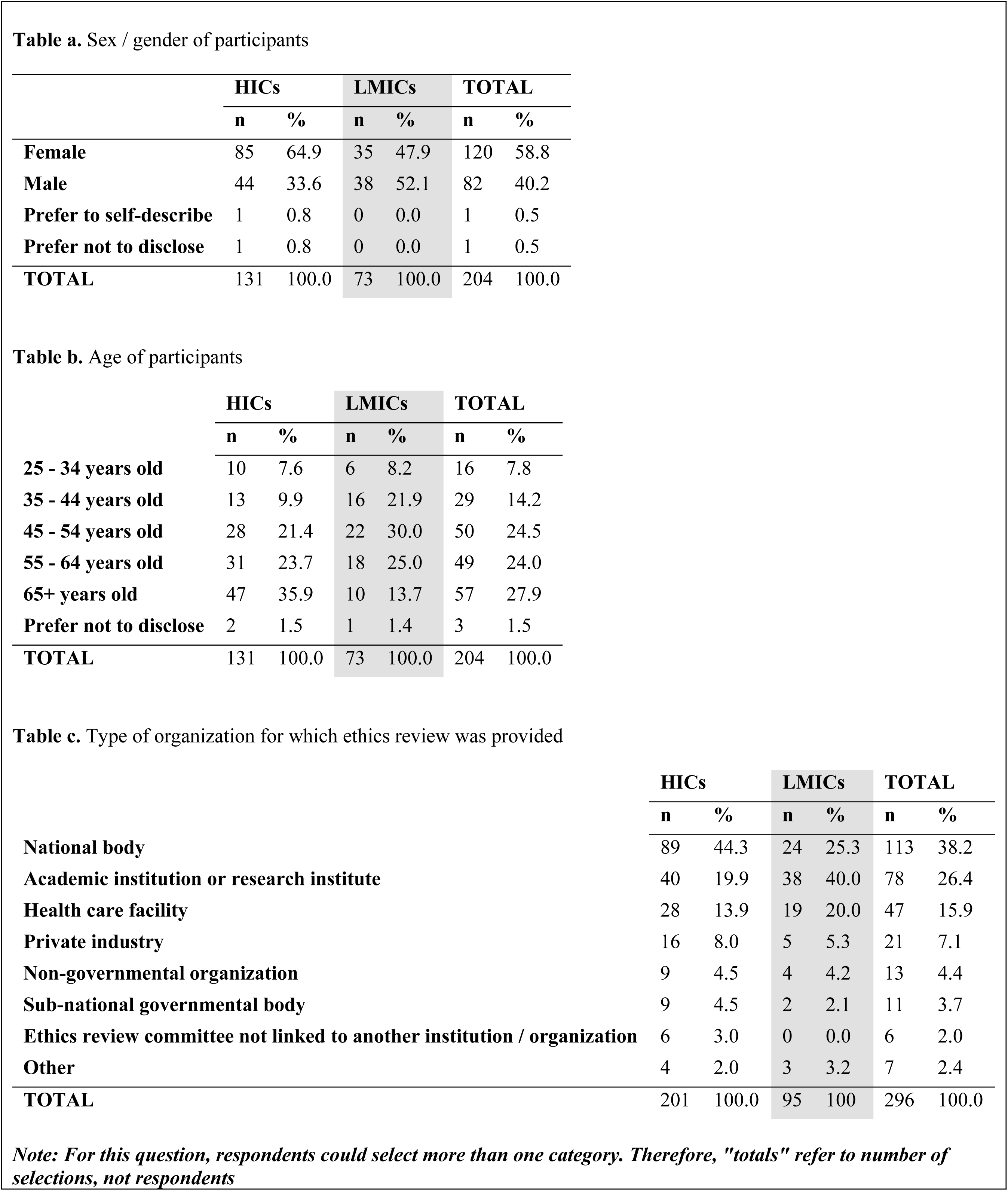

|  | HICs |  | LMICs |  | TOTAL |  |
| --- | --- | --- | --- | --- | --- | --- |
|  | n | % | n | % | n | % |
| **Female** | 85 | 64.9 | 35 | 47.9 | 120 | 58.8 |
| **Male** | 44 | 33.6 | 38 | 52.1 | 82 | 40.2 |
| **Prefer to self-describe** | 1 | 0.8 | 0 | 0.0 | 1 | 0.5 |
| **Prefer not to disclose** | 1 | 0.8 | 0 | 0.0 | 1 | 0.5 |
| **TOTAL** | 131 | 100.0 | 73 | 100.0 | 204 | 100.0 |

**Table b.** Age of participants
|  | HICs |  | LMICs |  | TOTAL |  |
| --- | --- | --- | --- | --- | --- | --- |
|  | n | % | n | % | n | % |
| **25 - 34 years old** | 10 | 7.6 | 6 | 8.2 | 16 | 7.8 |
| **35 - 44 years old** | 13 | 9.9 | 16 | 21.9 | 29 | 14.2 |
| **45 - 54 years old** | 28 | 21.4 | 22 | 30.0 | 50 | 24.5 |
| **55 - 64 years old** | 31 | 23.7 | 18 | 25.0 | 49 | 24.0 |
| **65+ years old** | 47 | 35.9 | 10 | 13.7 | 57 | 27.9 |
| **Prefer not to disclose** | 2 | 1.5 | 1 | 1.4 | 3 | 1.5 |
| **TOTAL** | 131 | 100.0 | 73 | 100.0 | 204 | 100.0 |

**Table c.** Type of organization for which ethics review was provided
|  | HICs |  | LMICs |  | TOTAL |  |
| --- | --- | --- | --- | --- | --- | --- |
|  | n | % | n | % | n | % |
| **National body** | 89 | 44.3 | 24 | 25.3 | 113 | 38.2 |
| **Academic institution or research institute** | 40 | 19.9 | 38 | 40.0 | 78 | 26.4 |
| **Health care facility** | 28 | 13.9 | 19 | 20.0 | 47 | 15.9 |
| **Private industry** | 16 | 8.0 | 5 | 5.3 | 21 | 7.1 |
| **Non-governmental organization** | 9 | 4.5 | 4 | 4.2 | 13 | 4.4 |
| **Sub-national governmental body** | 9 | 4.5 | 2 | 2.1 | 11 | 3.7 |
| **Ethics review committee not linked to another institution / organization** | 6 | 3.0 | 0 | 0.0 | 6 | 2.0 |
| **Other** | 4 | 2.0 | 3 | 3.2 | 7 | 2.4 |
| **TOTAL** | 201 | 100.0 | 95 | 100 | 296 | 100.0 |

*Note: For this question, respondents could select more than one category. Therefore, "totals" refer to number of selections, not respondents*

In terms of the WHO region for which ethics review was provided, all regions were represented in our survey (**Table S 1**). More than one third of respondents reviewed research protocols from Europe, almost one fifth from the Americas, one tenth from Africa, and less than one tenth each from the other WHO regions.

**Table 1** shows the number of respondents by country of residence. Participants from 48 countries (19 HICs and 29 LMICs) responded to our survey. Of the 203 individuals who indicated their country of residence, 130 (64%) were from HICs and 73 (36%) from LMICs. There was a large contingent of respondents from the UK (93).

**Fig. 1.**
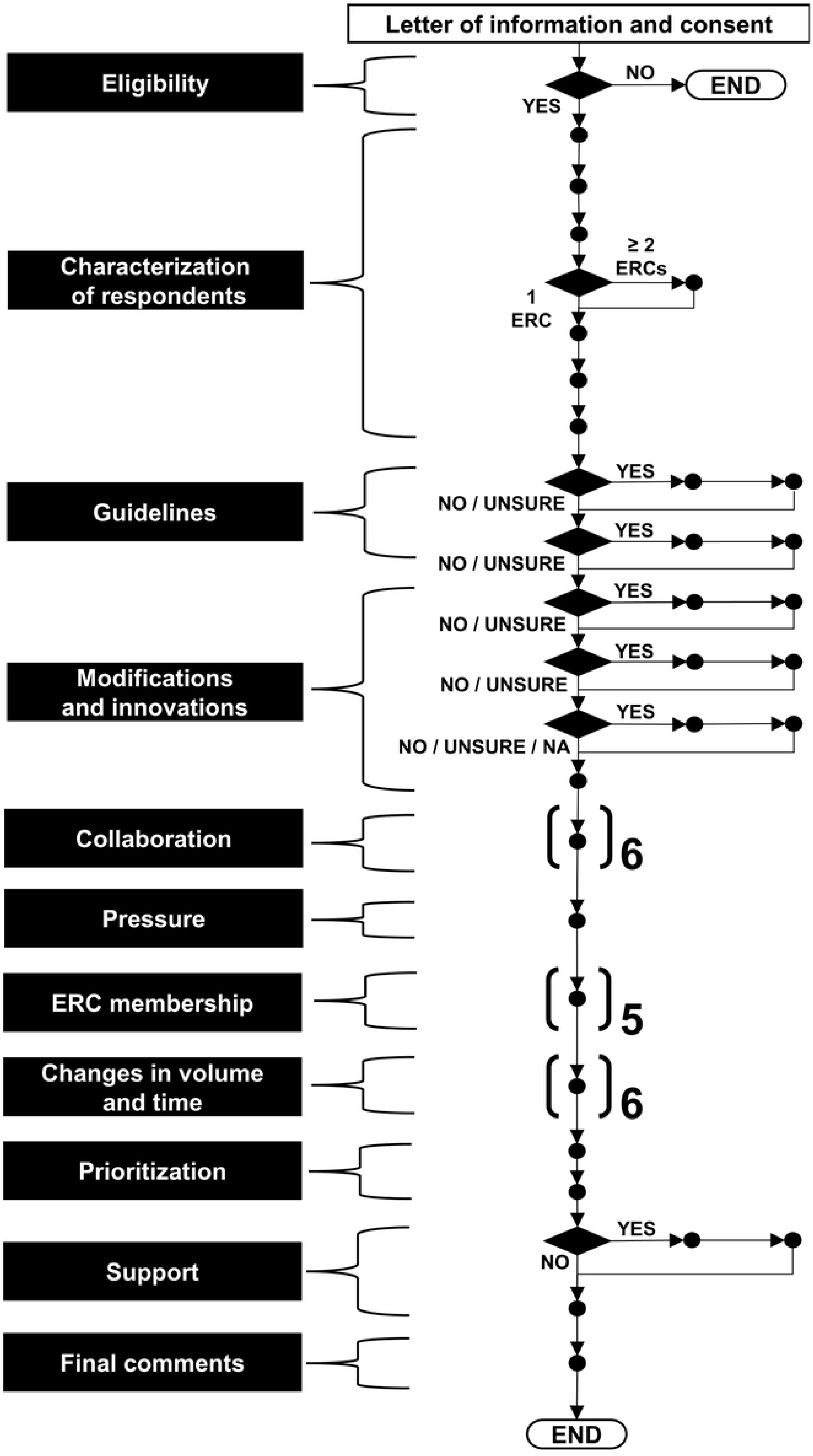
Overall structure and flow of the survey questionnaire

**Table 1.** Number of respondents by country of residence.

| Total number of countries: 48 |  |  |  |
| --- | --- | --- | --- |
| Total number of respondents who indicated country of residence: 203 |  |  |  |
| Respondents from 19 high-income countries (HICs) |  | Respondents from 29 low- and middle-income countries (LMICs) |  |
| Australia | 1 | Angola | 1 |
| Barbados | 1 | Argentina | 6 |
| Belgium | 2 | Bangladesh | 1 |
| Brunei Darussalam | 1 | Brazil | 4 |
| Canada | 3 | Cameroon | 1 |
| Chile | 2 | Colombia | 1 |
| Estonia | 1 | Costa Rica | 2 |
| France | 1 | Ecuador | 1 |
| Hungary | 1 | Egypt | 1 |
| Italy | 1 | El Salvador | 2 |
| Latvia | 1 | Ethiopia | 2 |
| New Zealand | 1 | Ghana | 1 |
| Panama | 1 | Grenada | 2 |
| Portugal | 5 | Honduras | 1 |
| San Marino | 1 | India | 6 |
| Singapore | 6 | Indonesia | 1 |
| South Korea | 2 | Iran | 2 |
| United Kingdom | 93 | Kenya | 3 |
| United States | 6 | Lebanon | 1 |
|  |  | Liberia | 1 |
|  |  | Malawi | 2 |
|  |  | Malaysia | 8 |
|  |  | Mexico | 8 |
|  |  | Morocco | 1 |
|  |  | Mozambique | 1 |
|  |  | Pakistan | 4 |
|  |  | Saint Vincent and the Grenadines | 1 |
|  |  | South Africa | 6 |
|  |  | Uganda | 2 |
| **TOTAL** | **130** | **TOTAL** | **73** |

Two thirds of respondents had six or more years of experience as ERC members. This is true for participants from both HICs and LMICs (**Table S 2**).

As shown in **Table S 3**, about one half of respondents (52%) were involved in only one ERC. This pattern was common for participants from HICs and LMICs. However, more than one third of respondents from HICs participated in three or more ERCs during the COVID-19 pandemic. Of those who indicated involvement with multiple ERCs, close to one half specified that such participation was simultaneous (**Table S 4**).

### Support for the operation of ERCs during the pandemic

As shown in **Table S 5**, an overwhelming majority (78.4%) of respondents indicated that their ERCs received no additional support for the operation of their committees during the pandemic. This lack of support was more pronounced in the case of ERCs in LMICs. For the minority of ERCs that did receive support, this consisted mainly of administrative and human resources, with one quarter of respondents from LMICs stating that their ERCs also received financial support, in contrast to only 12.5% of those from HICs (**Table S 6**). In terms of specific areas supported, participants from both HICs and LMICs mentioned teleconferencing and virtual meeting capabilities, information technology, support staff, assistance for ERC reviewers, and training of ERC members (**Table 2**). Interestingly, while 20% of respondents from HICs chose ERC support staff as one of the areas that received assistance, only 7.5% of those from LMICs did.

**Table 2.** Areas that received additional support during the COVID-19 pandemic.

|  | HICs |  | LMICs |  | TOTAL |  |
| --- | --- | --- | --- | --- | --- | --- |
|  | n | % | n | % | n | % |
| Teleconferencing and virtual meeting capabilities | 23 | 28.0 | 8 | 20.0 | 31 | 25.4 |
| Ethics committee support staff | 16 | 19.5 | 3 | 7.5 | 19 | 15.6 |
| Information technology (IT) support | 12 | 14.6 | 6 | 15.0 | 18 | 14.8 |
| Ethics committee reviewers | 10 | 12.2 | 6 | 15.0 | 16 | 13.1 |
| Training of ethics committee members | 9 | 11.0 | 6 | 15.0 | 15 | 12.3 |
| Other administrative costs | 4 | 4.9 | 5 | 12.5 | 9 | 7.4 |
| Security for ethics committee members | 5 | 6.1 | 1 | 2.5 | 6 | 4.9 |
| External experts | 1 | 1.2 | 4 | 10.0 | 5 | 4.1 |
| Data management and storage | 1 | 1.2 | 1 | 2.5 | 2 | 1.6 |
| Other | 1 | 1.2 | 0 | 0.0 | 1 | 0.8 |
| **TOTAL** | **82** | **100.0** | **40** | **100.0** | **122** | **100.0** |

*Note: For this question, respondents could select more than one category. Therefore, "totals" refer to number of selections, not respondents*

In their text answers, participants alluded to support for covering the costs of using online platforms for meetings and protocol review, and for acquiring or upgrading hardware such as laptops and webcams. In one ERC, members were able to claim costs of setting up teleconferencing and of telephone calls if dialling into a meeting. In other ERCs, information technology training was offered, along with technical support for the use of online platforms. It is important to note that almost half of respondents from HICs, but close to the totality (91.4%) of those from LMICs, indicated that their ERCs lacked any pre-pandemic financial planning that included provisions for the support of the committees during a public health emergency (**Table S 7**).

### Modification of existing procedures or policies

Respondents from both HICs and LMICs overwhelmingly (more than 75% of participants in both cases) reported that their ERCs modified existing procedures or policies to operate during the pandemic (**Table S 8**). The most frequently modified domain was meeting logistics, followed by meeting frequency and procedures for protocol review and approval (**Table S 9**).

In terms of modifications to review procedures, several participants pointed out in their text responses that their ERCs fast-tracked the review of pandemic-related studies, shortening the timeline to review and approve protocols. ERC members were expected to complete the review of these protocols within a few days and, in some cases, 24 hours. To facilitate such a quick turnaround, some ERCs created special sub-committees that would conduct very fast protocol review. Moreover, participants emphasized the importance of simplifying and increasing the flexibility of administrative processes. For example, several respondents indicated that their ERCs switched entirely to the use of online platforms for protocol review, eliminating the need for paper documents.

Numerous participants stated that all ERC meetings were conducted virtually (as opposed to face-to-face) during the pandemic, which, in their view, enabled ERC members and researchers to participate regardless of geographical location, prevented contagion, and allowed rapid turnaround of reviews.

Even in the case of virtual sessions, all other full meeting requirements such as quorum had to be met. Some ERCs modified their meetings to open a permanent slot in their agendas for COVID-19-related research or added urgent full meetings to discuss top-priority pandemic-related trial protocols. In other cases, members were permanently available to review COVID-19-related protocols, with those pertaining to other topics addressed less frequently.

While most respondents acknowledged the advantages of using online platforms during the pandemic to organize ERC meetings and to review research protocols, several participants highlighted the challenges that the use of such technologies entailed, particularly for new and more senior members of the ERCs who felt uncomfortable using these platforms. Some individuals deplored the loss of quality in the dynamics among ERC members (stilted conversations, fewer informal interactions) compared against the benefits of face-to-face meetings. Resistance to working online for some was compounded by difficulties accessing the internet and the lack of adequate electronic devices to do so.

Regarding the modification of protocol requirements, respondents mentioned the need to add safety procedures for study participants and members of the research teams, facilitating remote documentation of consent, and changing the policies regarding the use of non-anonymized data from health service and public health records for the duration of the pandemic to allow more unrestrained use of data. Some ERCs transitioned from requiring the physical signature of conflict-of-interest declaration forms to an email declaration.

As shown in **Table 3**, only a minority of respondents indicated that their ERCs conducted a formal evaluation of the success or failure of modifying existing procedures or policies (28% of participants from HICs and 17% of those from LMICs). More than one quarter of respondents did not know whether such modifications had been assessed.

**Table 3.** Evaluation of the success / failure of modifying existing procedures or policies.

|  | HICs |  | LMICs |  | TOTAL |  |
| --- | --- | --- | --- | --- | --- | --- |
|  | n | % | n | % | n | % |
| **Yes** | 24 | 27.9 | 8 | 17.0 | 32 | 24.1 |
| **No** | 36 | 41.9 | 29 | 61.7 | 65 | 48.9 |
| **Unsure** | 26 | 30.2 | 10 | 21.3 | 36 | 27.1 |
| **TOTAL** | 86 | 100.0 | 47 | 100.0 | 133 | 100.0 |

### Design and implementation of new procedures and policies

Almost two-thirds of respondents from both HICs and LMICs reported that their ERCs had designed and implemented new procedures and policies to address the challenges brought about by the pandemic (**Table S 10**). As in the case of modifications to ERC processes, innovations occurred mainly in the areas of meeting logistics and frequency, and procedures for protocol review and approval (**Table S 11**). This was the case for ERCs in both HICs and LMICs.

In their text responses, participants mentioned the development and implementation of new standard operating procedures (SOPs) and the integration of *ad hoc* committees, some including specialists, for urgent, accelerated protocol review. Such fast-track ERCs could review studies in one or two days, considerably shortening the time to complete reviews. One respondent considered the most successful innovation to be the formation of a “pool” of committee members ready to be convened at very short notice to quickly review COVID-19-related protocols. Such an *ad hoc* committee enabled applications to be reviewed and turned around very quickly.

The proportion of ERCs that formally evaluated the success or failure of new procedures and policies was analogous to that described for modifications to SOPs. **Table 4** shows that just 37% of respondents from HICs and 21% of those from LMICs reported that their ERCs conducted such an evaluation.

**Table 4.** Evaluation of the success / failure of designing and implementing new procedures or policies.

|  | HICs |  | LMICs |  | TOTAL |  |
| --- | --- | --- | --- | --- | --- | --- |
|  | n | % | n | % | n | % |
| **Yes** | 25 | 37.3 | 8 | 21.6 | 33 | 31.7 |
| **No** | 26 | 38.8 | 26 | 70.3 | 52 | 50.0 |
| **Unsure** | 16 | 23.9 | 3 | 8.1 | 19 | 18.3 |
| **TOTAL** | 67 | 100.0 | 37 | 100.0 | 104 | 100.0 |

### Permanently putting into effect modifications and innovations

A substantial majority of respondents (almost three quarters of those from HICs and more than four-fifths of those from LMICs) stated that many of the modifications and innovations to operating procedures implemented during the pandemic should be permanently put into effect (**Table S 12**), particularly in the areas of meeting logistics and frequency, procedures for protocol review and approval, and training of ethics review committee members in new or modified procedures (**Table S 13**). Several participants argued in their text responses that virtual online meetings should be a permanent feature of ERC operations, as they increase efficiency and preclude many of the disadvantages of face-to-face meetings. Another recommendation was to enable the integration of *ad hoc* committees during times of increased demand. Similarly, respondents emphasized the relevance of facilitating the incorporation of new expert members to the ERCs as required. However, 20% of participants from HICs and 50% of those from LMICs indicated that their ERCs had no support to permanently implement modifications or innovations established during the COVID-19 pandemic (**Table S 14**).

### Policies, procedures, and guidelines for public health emergencies

It is noteworthy that almost half of respondents from HICs and three-quarters of participants from LMICs indicated that their ERCs did not have internal policies, procedures, or guidelines before the pandemic that could orient members regarding the functioning of the committees during PHEs (**Table S 15**). Regarding the use of internal guidelines, some ERCs adapted existing documents, while others developed entirely new procedures. In the absence of specific internal guidelines, some SOPs explicitly privileged expedited review during health crises.

In contrast to the widespread absence of internal guidelines, the ERCs of one quarter of respondents from HICs and of almost half of those from LMICs used external guidelines not developed by their committees to govern their operation during the pandemic (**Table S 16**). Members of several committees referred to publicly available national and international guidelines. A selection of the most consulted documents appears in Box 2.

#### Box 2.

National and international external guidelines**\*** that survey respondents reported were used by their ERCs to manage operations during the COVID-19 pandemic

**International Health Organizations**

- Council for International Organizations of Medical Sciences, & World Health Organization (2016). International Ethical Guidelines for Health-related Research Involving Humans (Fourth Ed.). Council for International Organizations of Medical Sciences. https://doi.org/10.56759/rgxl7405
- Pan-American Health Organization (2020). Guidance for ethics oversight of COVID-19 research in response to emerging evidence. https://iris.paho.org/handle/10665.2/53021
- Pan-American Health Organization (2020). Guidance and strategies to streamline ethics review and oversight of COVID-19-related research. https://iris.paho.org/handle/10665.2/52089
- Pan-American Health Organization (2020). Template and operational guidance for the ethics review and oversight of COVID-19-related research. https://iris.paho.org/handle/10665.2/52086
- Pan-American Health Organization (2022). Catalyzing ethical research in emergencies. Ethics guidance, lessons learned from the COVID-19 pandemic, and pending agenda. https://iris.paho.org/handle/10665.2/56139
- Red de América Latina y el Caribe de Comités Nacionales de Bioética - United Nations Educational, Scientific and Cultural Organization (UNESCO) (2020). Ante las investigaciones biomédicas por la pandemia de enfermedad infecciosa por coronavirus Covid-19. https://redbioetica.com.ar/wp-content/uploads/2020/03/Declaracion-RED-ALAC-CNBS-Investigaciones-Covid-19.pdf
- World Health Organization (2016). Guidance for managing ethical issues in infectious disease outbreaks. World Health Organization. https://apps.who.int/iris/handle/10665/250580
- World Health Organization (2020). Key criteria for the ethical acceptability of COVID-19 human challenge studies. https://apps.who.int/iris/handle/10665/331976
- World Health Organization (2020). Guidance for research ethics committees for rapid review of research during public health emergencies. https://apps.who.int/iris/handle/10665/332206
- World Health Organization (2020). Ethical standards for research during public health emergencies: distilling existing guidance to support COVID-19 R&D. https://apps.who.int/iris/handle/10665/331507

**Bioethics centres**

- Nuffield Council of Bioethics (2020). Ethical considerations in responding to the COVID-19 pandemic. https://www.nuffieldbioethics.org/assets/pdfs/Ethical-considerations-in-responding-to-the-COVID-19-pandemic.pdf
- The Hastings Center: Berlinger N *et al.* (2020). Ethical Framework for Health Care Institutions Responding to Novel Coronavirus SARS-CoV-2 (COVID-19). Guidelines for Institutional Ethics Services Responding to COVID-19. https://www.thehastingscenter.org/ethicalframeworkcovid19/

**Scientific publications mentioned by respondents**

- Saxena et al. (2019). Ethics preparedness: facilitating ethics review during outbreaks - recommendations from an expert panel. https://bmcmedethics.biomedcentral.com/articles/10.1186/s12910-019-0366-x

**National guidelines**

*Argentina*

- Resolución 908/2020. Ministerio de Salud de Argentina: https://www.argentina.gob.ar/normativa/nacional/resoluci%C3%B3n-908-2020-337359/texto

*Brazil*

- Normativas da Comissão Nacional de Ética em Pesquisa: http://conselho.saude.gov.br/normativas-conep?view=default

*Costa Rica*

- Consejo Nacional de Investigación en Salud de Costa Rica (CONIS) (2020). COMUNICADO 2: Recomendaciones para realizar investigación biomédica durante el periodo de la emergencia sanitaria por COVID-19 en Costa Rica. https://www.ministeriodesalud.go.cr/gestores_en_salud/conis/circulares/comunicado_cec_oac_oic_20082020.pdf

*El Salvador*

- Comité Nacional de Ética de la Investigación en Salud de El Salvador (2015). Manual de procedimientos operativos estándar para comités de ética de la investigación en salud. https://www.cneis.org.sv/wp-content/uploads/2018/07/MANUAL-CNEIS.pdf

*India*

- Indian Council of Medical Research (2017). National ethical guidelines for biomedical and health research involving human participants. https://ethics.ncdirindia.org/asset/pdf/ICMR_National_Ethical_Guidelines.pdf
- Indian Council of Medical Research (2020).National guidelines for ethics committees reviewing biomedical & health research during COVID-19 pandemic. https://main.icmr.nic.in/sites/default/files/guidelines/EC_Guidance_COVID19_06_05_2020.pdf

*Kenya*

- Kenya Medical Research Institute Scientific and Ethics Review Unit (2019). KEMRI SERU guidelines for the conduct of research during the covid-19 pandemic in Kenya. https://www.kemri.go.ke/wp-content/uploads/2019/11/KEMRI-SERU_GUIDELINES-FOR-THE-CONDUCT-OF-RESEARCH-DURING-THE-COVID_8-June-2020_Final.pdf

*Malaysia*

- Garis Panduan Pengurusan COVID-19 di Malaysia No.5 [*COVID-19 Management Guidelines in Malaysia No.5*] (2020). Ministry of Health of Malaysia. https://covid-19.moh.gov.my/garis-panduan/garis-panduan-kkm

*Pakistan*

- Government of Pakistan National COVID Command and Operation Center (NCOC) Guidelines (2020). [No longer available, as NCOC ceased operations on April 1, 2022)]

*South Africa*

- Department of Health, Republic of South Africa (2015). Ethics in Health Research: Principles, Processes and Structures (2d Ed). https://www.sun.ac.za/english/research-innovation/Research-Development/Documents/Integrity%20and%20Ethics/DoH%202015%20Ethics%20in%20Health%20Research%20-%20Principles,%20Processes%20and%20Structures%202nd%20Ed.pdf

*South Korea*

- Government of the Republic of Korea (2014). Bioethics and Safety Act (Act No. 12844). https://elaw.klri.re.kr/eng_mobile/viewer.do?hseq=33442&type=part&key=36

*United Kingdom*

- United Kingdom Health Departments / Research Ethics Service (2022). Standard Operating Procedures for Research Ethics Committees (Version 7.6). https://www.hra.nhs.uk/documents/3090/RES_Standard_Operating_Procedures_Version_7.6_September_2022_Final.pdf. [In particular, several respondents from the UK mentioned Section 9 of this document, which addresses expedited review in situations such as public health emergencies.]
- Health Research Authority (2020). https://www.hra.nhs.uk/approvals-amendments/
- Health Research Authority (2020). https://www.hra.nhs.uk/covid-19-research/covid-19-guidance-sponsors-sites-and-researchers/
- Department of Health and Social Care (2020). Coronavirus (COVID-19): notification to organisations to share information. https://www.gov.uk/government/publications/coronavirus-covid-19-notification-of-data-controllers-to-share-information

*\*We defined “external guidelines” as those not developed internally by participants’ ERCs*

### Changes in workload

Respondents stated that the workload of ERC members increased considerably during the pandemic, both because of the increase in the number of protocols reviewed and due to the urgency that the approval of COVID-19-related studies demanded. More than half of participants indicated that the volume of protocols received for review increased, both for studies assigned to delegated / expedited review, and for protocols that underwent full review (**Table S 17**). The increase in the volume of protocols had unexpected consequences. For example, in one HIC, the number of applicants who were summoned to discuss their protocols with ERCs in online meetings increased proportionally to the escalation in the volume of protocols submitted. In another case, ERC members were burdened with additional tasks such as working closely with the investigators of rejected COVID-19 protocols to improve their applications until these could be approved.

In terms of the time it took ERCs to process and approve protocols during the pandemic, respondents confirmed in their text answers that the turnaround time for ERC review was markedly shortened, from weeks or even months to just a few days. In general, more than half of survey participants indicated that, before the pandemic, the duration of the review process, from the time of initial submission to full approval, was between three and eight weeks (**Table S 18**). In contrast, during the pandemic, this process was substantially reduced to less than two weeks for both delegated / expedited review and full review. However, this decrease was more pronounced in HICs than in LMICs (**Table S 19**). Unsurprisingly, the approval of COVID-19-related research protocols was faster than that of non-COVID-19 studies. More than two-thirds of respondents indicated that delegated / expedited review of COVID-19-related protocols took less than five weeks; this was the case for more than half of full reviews. The process was longer in LMICs, though (**Table S 20**). Conversely, protocol review was slightly longer for non-COVID-19 studies, except in the case of full reviews in LMICs, which participants reported took between three and more than 12 weeks (**Table S 21**).

### Presence of external pressure on ERCs

While only 14% of respondents from HICs reported that their ERCs were subjected to different types of external pressure to both approve and reject research protocols, one third of participants from LMICs (34%) faced such a challenge (**Table S 22**). The most frequent perceived demand involved pressures to rush studies through the review process at the expense of proper examination and ethical oversight. This was especially evident in the case of COVID-19 vaccine clinical trials. Some participants highlighted their defense of the autonomy of their ERCs in the face of external influences by using, for example, research policies developed and implemented specifically for the pandemic as a tool for transparent decision-making and as a safeguard against external pressures. One ERC successfully resisted government pressure to approve a research protocol related to a domestic PCR test, human trials of locally developed ventilators, and a placebo-controlled vaccine trial proposed despite the existence of six emergency-authorized vaccines and ongoing mass vaccination.

While some respondents acknowledged that entities such as national governments were understandably impatient for preventive, diagnostic, and therapeutic measures to combat the pandemic, they still emphasized the need for proper and thorough review of research protocols. One respondent stated that institutional authorities that favoured or sponsored certain studies sought their immediate approval and considered ERCs as inconvenient hindrances to achieve this goal. Several participants described instances in which ERCs, particularly in LMICs, received pressure to approve alternative medicine clinical trials.

### Types of COVID-19 protocols reviewed by ERCs

Given the range of challenges brought about by the COVID-19 pandemic, it was interesting to determine the proportion of protocols received by ERCs according to the research area in which they could be classified, namely, diagnostics, therapeutics, vaccines, pharmacovigilance, or other topics such as behavioural research. Our results suggest that between one-half and two-thirds of ERCs received from one to 10 studies in each area (**Table 5**). In other words, all areas of COVID-19 research were covered in these protocols in ERCs of both HICs and LMICs. However, it must be noted that between one-third and one-half of respondents could not classify the protocols received by their ERCs (perhaps due to not tracking such information).

**Table 5.** Number of COVID-19 protocols reviewed, by type of study.

|  | Diagnostics |  |  |  | Therapeutics |  |  |  | Vaccines |  |  |  | Pharmacovigilance |  |  |  | Other |  |  |  |
| --- | --- | --- | --- | --- | --- | --- | --- | --- | --- | --- | --- | --- | --- | --- | --- | --- | --- | --- | --- | --- |
|  | HICs |  | LMICs |  | HICs |  | LMICs |  | HICs |  | LMICs |  | HICs |  | LMICs |  | HICs |  | LMICs |  |
|  | n | % | n | % | n | % | n | % | n | % | n | % | n | % | n | % | n | % | n | % |
| **0** | 10 | 22.2 | 8 | 25.0 | 10 | 19.6 | 8 | 22.9 | 10 | 22.2 | 8 | 24.2 | 10 | 27.8 | 7 | 25.9 | 9 | 20.5 | 7 | 20.0 |
| **1 to 10** | 29 | 64.4 | 19 | 59.4 | 27 | 52.9 | 17 | 48.6 | 24 | 53.3 | 16 | 48.5 | 20 | 55.6 | 13 | 48.1 | 23 | 52.3 | 16 | 45.7 |
| **11 to 20** | 5 | 11.1 | 3 | 9.4 | 5 | 9.8 | 3 | 8.6 | 4 | 8.9 | 3 | 9.1 | 4 | 11.1 | 3 | 11.1 | 4 | 9.1 | 3 | 8.6 |
| **21 to 30** | 0 | 0.0 | 2 | 6.3 | 0 | 0.0 | 2 | 5.7 | 0 | 0.0 | 2 | 6.1 | 0 | 0.0 | 1 | 3.7 | 0 | 0.0 | 1 | 2.9 |
| **> 30** | 1 | 2.2 | 0 | 0.0 | 1 | 2.0 | 0 | 0.0 | 1 | 2.2 | 0 | 0.0 | 1 | 2.8 | 0 | 0.0 | 1 | 2.3 | 0 | 0.0 |
| **No response** | 0 | 0.0 | 0 | 0.0 | 8 | 15.7 | 5 | 14.3 | 6 | 13.3 | 4 | 12.1 | 1 | 2.8 | 3 | 11.1 | 7 | 15.9 | 8 | 22.9 |
| **TOTAL** | **45** | **100** | **32** | **100** | **51** | **100** | **35** | **100** | **45** | **100** | **33** | **100** | **36** | **100** | **27** | **100** | **44** | **100** | **35** | **100** |

### Prioritization of protocols for ethics review

Overwhelmingly, and as expected, participants reported that their ERCs considered COVID-19-related protocols urgent and thus prioritized their review and approval over that of others, particularly in terms of expediting the review of these studies and privileging their discussion during committee meetings. More than three quarters of respondents from HICs and almost two-thirds of those from LMICs indicated that their ERCs gave priority to COVID-19-related studies (**Table S 23**). In fact, in one case, an ERC stopped reviewing non-COVID-19-related protocols altogether. Some ERCs gave precedence to the review of COVID-19-related studies according to the priorities determined by their national governments. Others were assigned studies by an *ad hoc* national entity that triaged the research protocols. Interestingly, however, as shown in **Table S 23**, 15% of respondents from HICs and 27% of those from LMICs stated that their ERCs did not give priority to pandemic-related studies.

Furthermore, our results show that almost one-third of respondents from HICs and almost half of those from LMICs indicated that, for their ERCs, the review of some types of COVID-19-related studies took precedence over that of others (**Table S 24**). In their text responses, participants explicitly mentioned prioritizing clinical trials, particularly those focused on COVID-19 vaccine development and safety monitoring; studies related to therapeutic agents for the treatment of COVID-19; protocols about diagnostics and prognostic factors; epidemiological studies, including those related to the natural history of COVID-19 and serosurveillance; and research affecting public health policy. In the case of one ERC in a HIC with very low infection rates resulting from successful public health measures, priority was given to vaccine trials and observational research on vaccine monitoring and community incidence.

### Membership of ERCs during the pandemic

One of the main challenges that ERCs worldwide faced during the pandemic was making certain that the number and expertise of their members enabled the efficient operation of the committees under such demanding circumstances. Most survey respondents indicated that their ERCs were able to ensure quorum (80% of participants from HICs, but only 60% of those from LMICs); however, one-third of respondents from LMICs stated that quorum in their ERCs was infrequently met (**Table S 25**). Two-thirds of participants from HICs and three quarters of those from LMICs reported that their committees had taken measures to ensure continuity of adequate review of research protocols in case existing members became unavailable due to the pandemic (**Table S 26**).

ERCs in both HICs and LMICS did invite new members or appointed alternate ones to ensure quorum and inclusion of individuals with appropriate expertise. Yet, consulting expert non-members seems to have been preferred to incorporating individuals to the committee. Only 13% of respondents from HICs and 24% of those from LMICs indicated that their ERCs had added new members to accelerate protocol review during the pandemic (**Table S 27**). Similarly, 11% of participants from HICs and 37% of those from LMICs added new members with specific expertise (**Table S 28**). In contrast, almost one-third of individuals from HICs, but close to two-thirds of those from LMICs, stated that their committees had consulted expert non-members to address novel areas of research or provide enhanced scrutiny of research protocols (**Table S 29**). In their text responses, participants expressed that, in some cases, ERCs incorporated new members available at quick notice and who were comfortable with the use of online platforms for meetings and protocol review. A similar approach consisted of integrating virtual *ad hoc* committees solely to review COVID-19-related-protocols. For some ERCs, national legislation complicated getting additional support or adding new members. Another factor complicating the integration of ERCs was that clinical responsibilities of individuals directly in the care of COVID-19 patients soared, hindering their participation in committee meetings. One participant reported that some ERC members could not fulfill their duties in the ERC because they had become highly sought-after “media celebrity” experts.

Survey respondents suggested that it would be worthwhile to assess the psychological and emotional challenges that ERC members faced by having to evaluate protocols using new, unfamiliar procedures under extreme pressure. Also, it is worth reiterating that, according to several participants, many ERC members, particularly older ones, deplored the loss of features common to face-to-face meetings, such as a warmer, more informal and welcoming environment that favoured interpersonal interactions. Other respondents expressed their desire for constructive and supportive feedback and for more appreciative and generous gestures of gratitude for the extraordinary efforts of ERCs. However, a few participants considered that being able to respond in a useful way to a public health crisis as ERC members was very gratifying and validating.

### National and international collaboration

While ∼40% of respondents from HICs and LMICs reported the presence of national and international collaboration among ERCs to standardize emergency operations and procedures during the pandemic, almost one-third of participants from HICs were unsure about the existence of such collaboration (**Table S 30**). Almost half of respondents from HICs, but more than two-thirds of those from LMICs, indicated that their ERCs did not have strategies to harmonize multiple review processes (**Table S 31**).

Most participants (55% of those from HICs and 63% of those from LMICs) reported that their ERCs relied on established procedures to recognize and validate research protocol reviews conducted by other committees (**Table S 32**). About one half of respondents from HICs, but almost two-thirds of those from LMICs, affirmed that their ERCs collaborated with scientific committees that pre-reviewed or prioritized pandemic-related research protocols (**Table S 33**).

Almost 50% of participants from HICs, but little more than a third of those from LMICs, reported the presence of centralized ethics review of research protocols for multicentre studies related to COVID-19 (**Table S 34**). Conversely, one-third of respondents from HICs, but more than two-thirds of those from LMICs, stated that their ERCs did not consider the formation of Joint Scientific Advisory Committees, Data Safety Review Committees, Data Access Committees, or a Joint Ethics Review Committee with representatives of ethics committees of all institutions and countries involved in COVID-19-related research (**Table S 35**).

In their text responses, participants noted the need for better inter-ERC collaboration and communication at the national and international levels to share successful strategies and avoid effort duplication. A case of very successful national inter-ERC collaboration is worth mentioning. Respondents from one particular LMIC stated that, given the critical absence of the national entity responsible for health research ethics during the pandemic, ERCs throughout the country joined forces to create an *ad hoc* spontaneous informal national network of all ERC chairs and co-chairs (it also included members of the national drug regulator) to strengthen mutual support, enhance communication among ERCs, identify best practices, and share academic and ethics resources.

## DISCUSSION

ERCs faced considerable challenges during the COVID-19 pandemic. Demands were placed on them to urgently review an increased volume of protocols while maintaining rigour, all under suboptimal conditions and uncertainty. Yet, our findings suggest ERCs reviewed a greater volume of protocols and did so faster than before the pandemic. Against this backdrop, our results also reveal that, despite billions of dollars having been invested into the R&D ecosystem to support the COVID-19 research response, little to no additional resources were directed to ERCs to support and/or expedite their functions. This should be particularly sobering for those who raise complaints about ERCs being an “obstacle” to research [12–15]. It may also help to explain other challenges experienced by ERCs during the pandemic, such as the absence of internal policies or guidelines for adapting to a PHE, the collateral damage sustained from deprioritizing non-COVID-19 protocols, and the pressures felt to rush protocols through review.

Our finding that ERCs wish to sustain many of the modifications made to their operations during the COVID-19 pandemic should be interpreted in light of the fact that ERCs also report having received little or no support during the COVID-19 pandemic as well as exiguous support for the maintenance of any modifications they wish to make permanent into the future. If it is expected that our research ethics ecosystem learns from this experience and enhances operations for future threats, it is difficult to see how this will be possible without significant investment. While no one seems to disagree that the research ethics ecosystem should strive for greater efficiency and collaboration, especially during PHEs, investments are required to achieve these aims. Simply put, the experience of ERCs during the COVID-19 pandemic, while herculean in many respects, was a function of necessity and is unlikely to be sustainable.

Extant literature reporting the challenges faced by ERCs during the COVID-19 pandemic is scant and tends to be limited to the early phases of this PHE. Most studies published on this topic are confined to single countries or geographical regions, with only one study including 14 countries in Africa, Asia, Australia, and Europe[16]. Several of these contributions focus exclusively on one ERC, usually associated with an academic or health care institution. The literature includes descriptions of ERC operations during the pandemic in Central America and the Dominican Republic [17], China [18], Ecuador [19], Egypt [20], Germany [21], India [22–24], Iran [10], Ireland [25], Kenya [26], Kyrgyzstan [27], Latin America [28], the Netherlands [29], Pakistan [30], South Africa [31,32], Turkey [33], and the United States [34–36]. Most of these studies reported results from surveys, interviews, focus groups, and documentary analysis, including review of research protocols, ERC meeting minutes, and existing SOPs. Participants usually consisted of ERC chairpersons and members, clinical and biomedical researchers, institutional representatives, and laypeople. Most studies based on surveys and interviews included fewer than 30 respondents, with only some having more than 100 participants.

Our findings agree with this literature. Given that our study is truly global in scope, it considerably broadens what is known about the operation of ERCs during the COVID-19 pandemic and clears a path towards greater consensus on strategies to prepare for and respond during future PHEs.

In this literature, several studies emphasize the lack of support and resources to operate during the pandemic. The vast majority of ERCs made numerous modifications to their SOPs. In particular, the use of online platforms for ERC meetings and for protocol review was ubiquitous. However, ERC members across studies pointed out several disadvantages of such platforms, including lack of familiarity and technical know-how, particularly in the case of more senior members of the committees. Only a few institutions provided training, equipment, and technical support for the use of these online platforms. Consistent with our findings, almost no ERCs in these studies reported having internal policies, procedures, or guidelines to operate during a PHE. National regulations on this topic, where available, were often unclear, contradictory, rapidly changing, vague, or difficult to interpret. Conversely, several ERCs availed themselves of international guidelines (Box 2), in particular those prepared by WHO [37–40] and PAHO [41–44].

In terms of changes in workload, all ERCs in the studies mentioned earlier experienced a dramatic increase in the number of COVID-19-related protocols received, which had to be reviewed very quickly in the face of pressure from researchers, institutions, governments, and the media for expedited approvals. The surge in the volume of protocols, along with shortened timelines for turnaround, severely strained ERC members’ ability to conduct rigorous, thorough, high-quality assessments. Despite feeling overwhelmed, ERC members participating in these studies managed to fulfill their responsibilities, sometimes at great personal cost.

Given the urgency to examine and approve an ever-increasing number of COVID-19 research protocols, the studies report several strategies implemented by ERCs worldwide to prioritize their review. This frequently meant that the assessment of non-COVID-19-related studies was postponed or even abandoned. Similarly, non-interventional COVID-19 protocols were given secondary importance. Prioritization of COVID-19 protocols by type of study was rare.

Despite numerous staffing challenges, most ERCs in the studies examined were able to ensure quorum. In some cases, their institutions provided training sessions to update committee members on the rapidly changing landscape of basic and clinical knowledge about COVID-19. A less frequently used approach was to incorporate new members with relevant expertise into the ERCs. One common strategy across different countries was the integration of *ad hoc* committees focused exclusively on the review of COVID-19 -related protocols.

The topic of centralized review of pandemic-related research is rather contentious in this literature. While some studies report ERC members favouring such an approach, others consider that a single national ERC in charge of PHE-specific ethics review is bound to be unsuccessful due to the importance of local context in responding to PHEs. In Ecuador, forcing researchers to submit their protocols to a seven-member centralized *ad hoc* ERC caused considerable delays in the approval process and instead severely impeded the execution of COVID-19-related studies [19].

As shown in our results, in some countries ERCs strengthened collaboration networks during the pandemic. A notable case was the creation of a spontaneous, informal, *ad hoc* group in South Africa— the Research Ethics Support in COVID-19 Pandemic (RESCOP) —by ERC chairpersons and members as a response to the lack of national ethics guidance and the unexpected critical absence of the National Health Research Ethics Council at the most crucial moment in the pandemic [32]. This example highlights the clear need for national governance and oversight for research ethics to ensure accountability and responsiveness of ERCs [45].

A common topic of concern across ERCs in several countries was the set of unique challenges to obtaining informed consent during the pandemic, especially in the case of patients unable to give consent, such as those who were severely ill, isolated, or in the intensive care unit. Thus, it was necessary to find innovative alternative strategies to obtain consent.

## RECOMMENDATIONS

### Box 3.

Recommendations to strengthen the resiliency of ERCs during future public health emergencies.

- Increase and assign an adequate proportion of the budgets of ERCs for:

- Their continued operation during PHEs, especially in terms of online teleconferencing and review platforms
- Sustaining select modifications and innovations designed and implemented during PHEs
- Increase awareness of the value of ERCs in the research and development (R&D) ecosystem as a means of protecting research participants, ensuring social value, and promoting public trust in research outputs, rather than as a bureaucratic nuisance
- Evaluate the success or failure of modifications and innovations designed and implemented during PHEs
- Develop a “first aid kit” for each ERC that includes:

- Existing external guidelines for committee operation during a PHE
- Internal contingency plans designed by the ERC or its home institution that adapt existing external guidelines to local contexts
- A directory of expert non-members available for consultation
- Easy-to-follow checklists that incorporate the essential elements needed to function during a PHE
- Familiarize ERC members with the “first aid kit” through periodic capacity building activities
- Consider the psychological and emotional challenges that ERC members face during PHEs
- Devise strategies to defend and safeguard ERCs’ autonomy against external pressures
- Promote national and regional collaboration networks of ERCs that strengthen their resiliency during PHEs
- Facilitate collaboration between ERCs and scientific committees

## LIMITATIONS AND STRENGTHS

In terms of the limitations of our study, it would have been desirable to include participants from more countries, and a larger number of respondents from each country. It was probably difficult to reach a higher response rate due to “pandemic fatigue”. Non-native English speakers, especially in LMICs, may have excluded themselves from our survey. Absent or unreliable internet access could have limited the participation of some participants, particularly in LMICs. The number of ERC members that provided ethics review for health care facilities was relatively low. Despite the anonymity of their answers, respondents may have been reluctant to share specific instances of external pressures impinging upon their ERCs. The large number of participants from the UK (93 out of 281) likely skewed the results from HICs, and from experiences in the UK in particular.

We chose to present our results descriptively and did not perform any analytic tests for statistically significant differences in responses. This was because we were unable to determine a denominator, so we could not meet the requirements for many significance tests. Non-parametric tests could have been used, but we think reporting statistical significance in this context would not be informative. Non-response bias could also influence our results. This could be non-differential in its effects as our results cohere with the literature thus far reported.

To our knowledge, this is the first examination at a global level of the challenges faced by ERCs during the COVID-19 pandemic, and the strategies used to address them. Also, our study compares for the first time several dimensions of the operation of ERCs during the pandemic between committees in HICs and those in LMICs. All WHO regions were represented in our study, as participants from 48 countries (19 HICs and 29 LMICs) responded to our survey. There was an adequate balance in terms of the sex / gender of respondents. Furthermore, the ample experience of the study participants as ERC members (two thirds of respondents had six or more years of experience in this role) strengthens the generalizability of our findings. The recommendations suggested by the study participants are quite relevant to combating future public health emergencies. In general, all these strengths give credence to the validity, reliability, and accuracy of our results.

## Data Availability

All relevant data are within the manuscript and its Supporting Information files. Additionally, the raw survey data are available as two Excel spreadsheets from the Figshare database (DOI: 10.6084/m9.figshare.24076704)

https://doi.org/10.6084/m9.figshare.24076704

## ACKNOWLEDGEMENTS

We are very grateful to all the members of ethics review committees who participated in our survey. We also want to express our gratitude to Andreas Reis and Katherine Littler of the Health Ethics and Governance Unit, World Health Organization, and to the following members of the World Health Organization COVID-19 Ethics and Governance Working Group, for their inputs on the survey instrument and manuscript: Aasim Ahmad, Thalia Arawi, Caesar Atuire, Oumou Bah-Sow, Anant Bhan, Ingrid Callies, Angus Dawson, Jean-François Delfraissy, Ezekiel Emanuel, Ruth Faden, Tina Garanis-Papadatos, Prakash Ghimire, Dirceu Greco, Calvin Ho, Patrik Hummel, Zubairu Iliyasu, Mohga Kamal-Yanni, Sharon Kaur, So Yoon Kim, Sonali Kochhar, Ruipeng Lei, Ahmed Mandil, Julian März, Ignacio Mastroleo, Roli Mathur, Signe Mežinska, Ryoko Miyazaki-Krause, Keymanthri Moodley, Suerie Moon, Michael Parker, Carla Saenz, G. Owen Schaefer, Ehsan Shamsi-Gooshki, Jerome Singh, Beatriz Thomé, Teck Chuan Voo, Jonathan Wolff, and Xiaomei Zhai.

This project was funded by a Canadian Institutes of Health Research grant (#C150-2019-11).

## SUPPORTING INFORMATION

**S1 Appendix. Qualtrics questionnaire**

**S2 Appendix. Supplementary Tables**

- **Table S 1.** World Health Organization region for which ethics review was provided
- **Table S 2.** Length of experience of participants as ethics review committee members
- **Table S 3.** Number of ethics review committees in which participants were involved during the COVID-19 pandemic
- **Table S 4.** Simultaneous involvement with multiple ethics review committees
- **Table S 5.** Presence of additional support for the operation of ethics review committee during the COVID-19 pandemic
- **Table S 6.** Additional support of ethics review committee, by type
- **Table S 7.** Pre-pandemic financial planning with provisions for support of ethics review committees during a public health emergency
- **Table S 8.** Modification of existing procedures or policies
- **Table S 9.** Modified procedures and policies
- **Table S 10.** Design and implementation of new procedures and policies
- **Table S 11.** New procedures and policies
- **Table S 12.** Opinions on whether to permanently put into effect modifications or innovations to operating procedures implemented during the COVID-19 pandemic
- **Table S 13.** Modifications or innovations to operating procedures implemented during the COVID-19 pandemic that should be permanently put into effect
- **Table S 14.** Presence of support to permanently implement modifications or innovations established during the COVID-19 pandemic
- **Table S 15.** Presence of internal policies, procedures, or guidelines
- **Table S 16.** Presence of external policies, procedures, or guidelines
- **Table S 17.** Extent of change in the volume of protocols reviewed during the pandemic
- **Table S 18.** Time it took before the COVID-19 pandemic for research protocols to be approved, from the time of initial submission to full approval
- **Table S 19.** Length of time that ethics review committee members took to complete review of research protocols during the COVID-19 pandemic
- **Table S 20.** Total time to approval of COVID-19-related research protocols
- **Table S 21.** Total time to approval for non-COVID-19-related research protocols
- **Table S 22.** Presence of external pressure on ethics review committees to approve or reject specific COVID-19 research protocols
- **Table S 23.** Prioritization of COVID-19 related research over non-COVID-19-related protocols
- **Table S 24.** Prioritization of some types of COVID-19-related research over others
- **Table S 25.** Ensuring quorum
- **Table S 26.** Presence of measures to ensure continuity of adequate review of research protocols in case existing members became unavailable due to the pandemic
- **Table S 27.** Addition of new members to accelerate protocol review during the COVID-19 pandemic
- **Table S 28.** Addition of new members with specific expertise to address novel areas of research or provide enhanced scrutiny of research protocols during the COVID-19 pandemic
- **Table S 29.** Consultation of expert non-members to address novel areas of research or provide enhanced scrutiny of research protocols during the COVID-19 pandemic
- **Table S 30.** National and international collaboration among ethics review committees to standardize emergency operations and procedures during the COVID-19 pandemic
- **Table S 31.** Presence of strategies to harmonize multiple review processes
- **Table S 32.** Reliance on established procedures to recognize and validate research protocol reviews conducted by other ethics committees
- **Table S 33.** Collaboration with scientific committees that pre-reviewed or prioritized pandemic-related research protocols
- **Table S 34.** Presence of centralized ethics review of research protocols for multicentre studies related to COVID-19
- **Table S 35.** Formation of Joint Scientific Advisory Committees, Data Safety Review Committees, Data Access Committees, or a Joint Ethics Review Committee

## REFERENCES

1. Aarons D. Addressing the challenge for expedient ethical review of research in disasters and disease outbreaks. Bioethics. 2019;33: 343–346. doi:10.1111/bioe.12543

2. Alirol E, Kuesel AC, Guraiib MM, De la Fuente-Núñez V, Saxena A, Gomes MF. Ethics review of studies during public health emergencies - The experience of the WHO ethics review committee during the Ebola virus disease epidemic. BMC Med Ethics. 2017;18: 1–12. doi:10.1186/s12910-017-0201-1

3. Council for International Organizations of Medical Sciences, World Health Organization. International Ethical Guidelines for Health-related Research Involving Humans. Fourth Ed. Geneva: Council for International Organizations of Medical Sciences; 2016.

4. Schopper D, Upshur R, Matthys F, Singh JA, Bandewar SS, Ahmad A, et al. Research ethics review in humanitarian contexts: The experience of the independent ethics review board of Médecins Sans Frontières. PLoS Med. 2009;6: 1–6. doi:10.1371/journal.pmed.1000115

5. Schopper D, Ravinetto R, Schwartz L, Kamaara E, Sheel S, Segelid MJ, et al. Research ethics governance in times of Ebola. Public Health Ethics. 2017;10: 49–61. doi:10.1093/phe/phw039

6. Tansey CM, Herridge MS, Heslegrave RJ, Lavery J V. A framework for research ethics review during public emergencies. Can Med Assoc J. 2010;182: 1533–1537. doi:10.1503/cmaj.090976

7. Canadian Institutes of Health Research, Natural Sciences and Engineering Council of Canada, Social Sciences and Humanities Research Council. Tri-Council Policy Statement: Ethical Conduct for Research Involving Humans. 2018. Available: https://ethics.gc.ca/eng/tcps2-eptc2_2018_chapter6-chapitre6.html#d

8. London AJ, Kimmelman J. Against pandemic research exceptionalism. Science (1979). 2020;368: 476–477. doi:10.1126/science.abc1731

9. Moodley K, Hardie K, Selgelid MJ, Waldman RJ, Strebel P, Rees H, et al. Ethical considerations for vaccination programmes in acute humanitarian emergencies. Bull World Health Organ. 2013;91: 290–297. doi:10.2471/BLT.12.113480

10. Hashemi A, Bahmani F, Tehrani SS, Forouzandeh M, Koohpayehzadeh J, Ashrafi M, et al. Ethical considerations and interdisciplinary approach to research on COVID-19 pandemic: The response of Iran University of Medical Sciences. Med J Islam Repub Iran. 2020;34: 1–4. doi:10.34171/mjiri.34.87

11. Lencucha R, Neupane S. The use, misuse and overuse of the “low-income and middle-income countries” category. BMJ Glob Health. 2022;7. doi:10.1136/bmjgh-2022-009067

12. Schneider CE. The Censor’s Hand: The Misregulation of Human-Subject Research. 1st edition. Cambridge, Massachusetts: The MIT Press; 2015.

13. Kotsis S V., Chung KC. Institutional review boards: What’s old? what’s new? what needs to change? Plast Reconstr Surg. 2014;133: 439–445. doi:10.1097/01.prs.0000436846.00247.73

14. Stryjewski TP, Kalish BT, Silverman B, Lehmann LS. The Impact of Institutional Review Boards (IRBs) on clinical innovation: A survey of investigators and IRB members. J Empir Res Hum Res Ethics. 2015;10: 481–487. doi:10.1177/1556264615614936

15. Whitney SN. Institutional review boards: A flawed system of risk management. Res Ethics. 2016;12: 182–200. doi:10.1177/1747016116649993

16. Bauer A, Eskat A, Ntekim A, Wong C, Eberle D, Hedayati E, et al. How COVID-19 changed clinical research strategies: a global survey. J Int Med Res. 2022;50. doi:10.1177/03000605221093179

17. Canario Guzmán JA, Orlich J, Mendizábal-Cabrera R, Ying A, Vergès C, Espinoza E, et al. Strengthening research ethics governance and regulatory oversight in Central America and the Dominican Republic in response to the COVID-19 pandemic: a qualitative study. Health Res Policy Syst. 2022;20. doi:10.1186/s12961-022-00933-z

18. Zhang H, Shao F, Gu J, Li L, Wang Y. Ethics Committee Reviews of Applications for Research Studies at 1 Hospital in China During the 2019 Novel Coronavirus Epidemic. JAMA. 2020;323: 1844–1846. doi:10.1001/jama.2020.3786

19. Sisa I, Mena B, Teran E. The negative impact of ad hoc committees for ethical evaluation: The case of COVID-19-related research in Ecuador. Dev World Bioeth. 2021;21: 3–6. doi:10.1111/dewb.12307

20. Marzouk D, Sharawy I, Nakhla I, El Hodhod M, Gadallah H, El-Shalakany A, et al. Challenges During Review of COVID-19 Research Proposals: Experience of Faculty of Medicine, Ain Shams University Research Ethics Committee, Egypt. Front Med (Lausanne). 2021;8: 715796– 715804. doi:10.3389/fmed.2021.715796

21. Faust A, Sierawska A, Krüger K, Wisgalla A, Hasford J, Strech D. Challenges and proposed solutions in making clinical research on COVID-19 ethical: a status quo analysis across German research ethics committees. BMC Med Ethics. 2021;22. doi:10.1186/s12910-021-00666-8

22. Joshi SG, Safai AA, Barve SS. Experience of the selected Ethics Committee of Pune city regarding the review of COVID-19 protocols during the pandemic. Perspect Clin Res. 2023;14: 43–44. doi:10.4103/picr.picr_2_22

23. Kadam A, Patil S, Sane S, Shahabuddin S, Panda S. Challenges faced by ethics committee members in India during COVID-19 pandemic: A mixed-methods exploration. Indian J Med Res. 2022;155: 461–471. doi:10.4103/ijmr.ijmr_1095_22

24. Mukherjee S, Samajdar S, Tripathi R, Tripathi S. Functioning of Institutional Ethics Committees during the COVID-19 pandemic: An opinion survey. Perspect Clin Res. 2022;13: 118–119. doi:10.4103/picr.PICR_103_21

25. Sheehy A, Ralph James J, Horgan M. Implementing a National Approach to Research Ethics Review during a Pandemic – the Irish Experience. HRB Open Res. 2020;3: 63. doi:10.12688/hrbopenres.13146.1

26. Hinga A, Jeena L, Awuor E, Kahindi J, Munene M, Kinyanjui S, et al. Pandemic preparedness and responsiveness of research review committees: lessons from review of COVID-19 protocols at KEMRI Wellcome Trust Research Programme in Kenya. Wellcome Open Res. 2022;7: 75. doi:10.12688/wellcomeopenres.17533.1

27. Kudaibergenova T, Ibrahim M, Jain N, Vetra J. Documentary Assessment of the Abilities of Kyrgyzstan’s Research Ethics Committees During Public Health Emergency and Non-Emergency Situations. J Empir Res Hum Res Ethics. 2023;18: 99–108. doi:10.1177/15562646231176711

28. Palmero A, Carracedo S, Cabrera N, Bianchini A. Governance frameworks for COVID-19 research ethics review and oversight in Latin America: an exploratory study. BMC Med Ethics. 2021;22: 147–156. doi:10.1186/s12910-021-00715-2

29. Ijkema R, Janssens MJPA, van der Post JAM, Licht CM. Ethical review of COVID-19 research in the Netherlands; a mixed-method evaluation among medical research ethics committees and investigators. PLoS One. 2021;16: e0255040. doi:10.1371/journal.pone.0255040

30. Shekhani S, Iqbal S, Jafarey A. Adapting the ethical review process for COVID-19 research: reviewers’ perspectives from Pakistan. East Mediterr Health J. 2021;27: 1045–1051. doi:10.26719/emhj.21.053

31. Burgess T, Rennie S, Moodley K. Key ethical issues encountered during COVID-19 research: a thematic analysis of perspectives from South African research ethics committees. BMC Med Ethics. 2023;24: 11–24. doi:10.1186/s12910-023-00888-y

32. Rossouw TM, Wassenaar D, Kruger M, Blockman M, Hunter A, Burgess T. Research ethics support during the COVID-19 epidemic: a collaborative effort by South African Research Ethics Committees. S Afr Health Rev. 2021;2021: 163–172.

33. Ekmekci PE, Güner MD, Buruk B, Güneş B, Arda B, Görkey Ş. Challenges and practices arising during public health emergencies: A qualitative survey on ethics committees. Dev World Bioeth. 2023;23: 23–33. doi:10.1111/dewb.12345

34. Ford DE, Johnson A, Nichols JJ, Rothwell E, Dubinett S, Naeim A. Challenges and lessons learned for institutional review board procedures during the COVID-19 pandemic. J Clin Transl Sci. 2021;5: e107. doi:10.1017/cts.2021.27

35. Sisk BA, Baldwin K, Parsons M, DuBois JM. Ethical, regulatory, and practical barriers to COVID-19 research: A stakeholder-informed inventory of concerns. PLoS One. 2022;17: e0265252. doi:10.1371/journal.pone.0265252

36. Taylor HA, Serpico K, Lynch HF, Baumann J, Anderson EE. A snapshot of U.S. IRB review of COVID-19 research in the early pandemic. J Clin Transl Sci. 2021;5: e205. doi:10.1017/cts.2021.848

37. World Health Organization. Guidance for managing ethical issues in infectious disease outbreaks. Geneva; 2016. Available: https://apps.who.int/iris/handle/10665/250580

38. World Health Organization. Key criteria for the ethical acceptability of COVID-19 human challenge studies 6 May 2020. 2020. Available: https://apps.who.int/iris/handle/10665/331976

39. World Health Organization. Guidance for research ethics committees for rapid review of research during public health emergencies. 2020. Available: https://apps.who.int/iris/handle/10665/332206

40. World Health Organization. Ethical standards for research during public health emergencies: Distilling existing guidance to support COVID-19 R&D. Journal of Infectious Diseases. 2020. Available: https://apps.who.int/iris/handle/10665/331507

41. Pan-American Health Organization. Catalyzing ethical research in emergencies: Ethics guidance, lessons learned from the COVID-19 pandemic, and pending agenda. 2022. Available: https://iris.paho.org/handle/10665.2/56139

42. Pan-American Health Organization. Template and operational guidance for the ethics review and oversight of COVID-19-related research. 2020. Available: https://iris.paho.org/handle/10665.2/52086

43. Pan-American Health Organization. Guidance and strategies to streamline ethics review and oversight of COVID-19-related research. 2020. Available: https://iris.paho.org/handle/10665.2/52089

44. Pan-American Health Organization. Guidance for ethics oversight of COVID-19 research in response to emerging evidence. 2020. Available: https://iris.paho.org/handle/10665.2/53021

45. World Health Organization. WHO tool for benchmarking ethics oversight of health-related research with human participants (Draft). Geneva; 2022. Available: https://www.who.int/publications/m/item/who-tool-for-benchmarking-ethics-oversight-of-health-related-research-with-human-participants

